# Vascular Encasement Image Defined Risk Factors Predict Surgical Complications in Neuroblastoma

**DOI:** 10.1101/2022.07.05.22277282

**Authors:** Rachael Stokes, Aidan Bannon, Bonnie Leung, Jasmin Alloo, David Davies-Payne, Mark Winstanley, Andrew Wood, Stephen Evans, James Hamill

## Abstract

**Background:** Specific Image Defined Risk Factors (IDRF) may be of more relevance to the pediatric surgical oncologist than simply the presence of any IDRF. The aim of this study was to correlate IDRF with surgical complications with reference to vascular encasement IDRF and the grade of complication.

**Methods:** We searched the New Zealand Children’s Cancer Registry for all cases of neuroblastoma treated at a single pediatric oncology center between January 2007 – February 2021 and reviewed the pre-treatment axial imaging for IDRF status. Surgical complications were scored by Clavien-Dindo grade and correlated with the number of IDRF and with the subset of vascular encasement IDRF.

**Results:** Of 101 patients, 77 were IDRF positive. In total, 74 underwent surgical resection and 32 (43.2%) had a surgical complication. Complications were related to the number of IDRF (OR 1.33, 95% CI 1.05 – 1.73, p = 0.02) and the subgroup of vascular encasement IDRF (OR 1.78, 95% CI 1.12 – 3.04, p = 0.01) but were not significantly correlated with the subgroup of non-vascular encasement IDRF. We report three cases of chyle leak associated with tumor encasing the origin of the celiac axis and/or the superior mesenteric artery.

**Conclusions:** The vascular encasement IDRF subgroup is potentially a more useful prognostic indicator of surgical complications than non-vascular IDRF. More studies are needed to correlate specific IDRF with specific surgical complications to aid operative decision making.

**Level of evidence:** Level III

**Highlights:**

- Image Defined Risk Factors (IDRF) in neuroblastoma correlate with survival, completeness of resection, and surgical complications.
- Complications correlate specifically with the subset of vascular encasement IDRF. Chylous ascites was a specific complication associated with encasement of the celiac and superior mesenteric vessels.

## Introduction

Image Defined Risk Factors (IDRF) play a key role in the staging and risk classification of neuroblastoma. The IDRF system evolved from ‘surgical risk factors’ used by the European International Society of Pediatric Oncology Neuroblastoma Group. Cecchetto et al. showed that surgical risk factors increased the rate of postoperative complications and decreased the chance of complete surgical resection [1]. Surgical risk factors were renamed Image Defined Risk Factors and incorporated into the International Neuroblastoma Risk Group Staging System (INRGSS) which consists of four stages: L1, L2, M, and MS, where stage L1 represents localized tumor with no IDRF present and Stage L2 represents localized neuroblastoma with one or more IDRF [2]. The INRGSS in turn contributes to the International Neuroblastoma Risk Group system in which the patient’s age, tumor stage, histology, and genomic biomarkers (for example, *MYCN* status) categorize patients into four risk groups: very low-risk, low-risk, intermediate-risk, and high-risk. The neuroblastoma risk group system was revised in 2021 to incorporate single chromosome aberration status [3]. Therefore, IDRF contributes to the oncological risk classification of children with neuroblastoma.

IDRF status influences surgical planning, for example, tumors with no IDRF can be resected up front while IDRF positive tumors usually receive neoadjuvant chemotherapy followed by surgery. Completeness of resection in high-risk tumors is important because complete or near-complete surgical resection has been shown to improve survival [4–6]. Completeness of resection is partially determined by the number and type of IDRF present prior to surgery [7]. IDRF status also influences the chances of a complication after the operation [6,8]. Of emerging interest is how specific types of IDRF, not just the presence or absence of an IDRF, influence surgical complications. For example, Temple et al. showed that vascular encasement IDRF, where tumor surrounds a major blood vessel by more than 50%, are a subgroup of IDRF that correlate with more complications [9]. This, however, was not confirmed by van Heerden et al. who found the vascular encasement type of IDRF was not significantly associated with complications but organ invasion and tumor in two body compartments were [10]. This shows that the picture is not clear and, as noted by Cecchetto et al., more work needs to be done on how specific groups of IDRF increase the risk of specific complications [1]. Therefore, we reviewed our experience with neuroblastoma with respect to IDRF and surgical complications.

The purpose of the present study was to confirm reports on the correlation between the number and type of IDRF and surgical complications, and to report our experience with specific complications and their association with specific IDRF.

## Methods

This study was registered with the ADHB Research Review Committee, number A+8215. We searched the New Zealand Children’s Cancer Registry for all cases of neuroblastoma treated at our pediatric oncology center between January 2007 and February 2021. Our center is a tertiary referral children’s hospital and one of two pediatric oncology centers in New Zealand. Over the study period, two pediatric surgeons performed the majority of neuroblastoma resections.

Data collected included gender, age at diagnosis, tumor site, pre-treatment risk category, INRGSS stage, histology, differentiation, *MYC-N, CHD-5*, type of surgical procedure, and completeness of resection (percentage determined by the surgeon intra-operatively).

Radiological assessment of IDRF was performed by a pediatric radiology fellow (BL) who reviewed all preoperative CT and MR images of the chest, abdomen, and pelvis and classified IDRF using published guidelines [11,12]. We classified eight IDRF in the vascular encasement type as shown in Table 1.

**Table 1.** Image Defined Risk Factors (IDRF) with vascular encasement IDRF noted.

| **Tumor site** | **IDRF** | **#** | **VE** |
| --- | --- | --- | --- |
| Ipsilateral tumor extension within two body compartments | Neck-chest, chest-abdomen, abdomen-pelvis | 1 |  |
| Neck | Tumor encasing carotid and/or vertebral artery and/or internal jugular vein | 2 | ✓ |
|  | Tumor extending to base of skull | 3 |  |
|  | Tumor compressing the trachea | 4 |  |
| Cervico-thoracic junction | Tumor encasing brachial plexus roots | 5 |  |
|  | Tumor encasing subclavian vessels and/or vertebral and/or carotid artery | 6 | ✓ |
|  | Tumor compressing the trachea | 7 |  |
| Thorax | Tumor encasing the aorta and/or major branches | 8 | ✓ |
|  | Tumor compressing the trachea and/or principal bronchi | 9 |  |
|  | Lower mediastinal tumor, infiltrating the costo-vertebral junction between T9 and T12 | 10 |  |
| Thoraco-abdominal | Tumor encasing the aorta and/or vena cava | 11 | ✓ |
| Abdomen/pelvis | Tumor infiltrating the porta hepatis and/or the hepatoduodenal ligament | 12 |  |
|  | Tumor encasing branches of the superior mesenteric artery at the mesenteric root | 13 | ✓ |
|  | Tumor encasing the origin of the celiac axis, and/or of the superior mesenteric artery | 14 | ✓ |
|  | Tumor invading one or both renal pedicles | 15 |  |
|  | Tumor encasing the aorta and/or vena cava | 16 | ✓ |
|  | Tumor encasing the iliac vessels | 17 | ✓ |
|  | Pelvic tumor crossing the sciatic notch | 18 |  |
| Intraspinal tumor extension whatever the location provided that: | More than one third of the spinal canal in the axial plane is invaded and/ or the perimedullary leptomeningeal spaces are not visible and/or the spinal cord signal is abnormal | 19 |  |
| Infiltration of adjacent organs/structures | Pericardium, diaphragm, kidney, liver, duodeno-pancreatic block, and mesentery | 20 |  |
| Conditions to be recorded, but not considered IDRF | Multifocal primary tumors |  |  |
|  | Pleural effusion, with or without malignant cells |  |  |
|  | Ascites, with or without malignant cells |  |  |

IDRF, image defined risk factors. VE, vascular encasement. Source, [2].

Complications included those documented in the operation note, discharge summary, or clinic letters. These were classified according to the Clavien-Dindo grade as follows: Grade I - deviation from normal course without the need for pharmacological, surgical, endoscopic or radiological interventions; Grade II – requires pharmacological treatment with drugs, including blood transfusions and total parenteral nutrition; Grade III – requiring surgical, endoscopic, or radiological intervention; Grade IV – life-threatening requiring intensive care unit management; and Grade V – death [13].

Statistical analysis was performed in R [14] using the packages Tidyverse [15] and Survival [16]. The distribution of continuous variables was tested for normality by the Shapiro-Wilk test (all were found to be nonparametric). Univariate analysis was performed using the Wilcoxon rank sum test and Fisher’s exact test. Multivariate analysis of factors that contributed to the occurrence of a complication was performed in a generalized linear model with a binomial fit. The outcome variable was the occurrence of a surgical complication (binomial), and explanatory variables were the number of IDRF, patient’s age, tumor site, histology, *MYC-N*, and *CHD-5*. Analysis of the effect of IDRF on the grade of complication, in those patients who developed a complication, was performed in a cumulative link model using the package, ordinal [17]. The outcome was the Clavien-Dindo grade of complication (as an ordinal factor), and explanatory variables were the number of IDRF, patient’s age, tumor site, histology, *MYC-N*, and *CHD-5*. P-values were obtained by comparison with a null model using analysis of variance (anova). Results are reported as median and interquartile range (IQR). Effect size from the models is reported as odds ratio (OR) and 95% confidence interval (95% CI).

## Results

### Patient and tumor characteristics

The registry identified 101 children with neuroblastoma between 2007 and 2021. The median age at diagnosis was 23 months (range 0 – 175months); 55 were 18 months of age or older. Patient and tumor characteristics by the presence or absence of IDRF are shown in Table 2.

**Table 2.** Baseline characteristics of all patients. Univariate analysis of the 74 patients who underwent a surgical resection compares those who had an uncomplicated postoperative course with those who experienced a complication.

|  | All patients*<br>n = 101 | Surgical resection patients<br>n = 73 |  | P-value** |
| --- | --- | --- | --- | --- |
|  |  | No surgical<br>complication<br>n = 37 | Surgical<br>complication<br>n = 36 |  |
| **Age** |  |  |  |  |
| Median months<br>[IQR]) | 23 [9, 42] | 16 [8, 34] | 35 [13.75, 53] | 0.01 |
| **Gender** |  |  |  |  |
| Female | 49 | 17 | 20 | 0.5 |
| Male | 52 | 20 | 16 |  |
| **Ethnicity** |  |  |  |  |
| Māori | 21 | 7 | 7 | 0.9 |
| Pacific | 10 | 4 | 2 |  |
| European | 61 | 21 | 23 |  |
| Other | 9 | 5 | 4 |  |
| **Site** |  |  |  |  |
| Abdominal/Pelvic | 75 | 27 | 25 | 0.3 |
| Thoracic | 21 | 8 | 11 |  |
| Neck | 5 | 2 | 0 |  |
| **Side** |  |  |  |  |
| Left | 47 | 18 | 17 | 0.5 |
| Right | 39 | 16 | 13 |  |
| Midline | 14 | 3 | 6 |  |
| **IDRF present** |  |  |  |  |
| No | 23 | 12 | 6 | 0.2 |
| Yes | 77 | 26 | 30 |  |
| **Number of IDRF** |  |  |  |  |
| Median [IQR] | 2 [1, 5] | 2 [0, 3] | 4 [1, 6] | 0.005 |
| **Number of VE IDRF** |  |  |  |  |
| Median [IQR] | 0 [0, 2] | 0 [0, 1] | 1 [0, 3] | 0.01 |
| **INRG Stage** |  |  |  |  |
| L1 | 20 | 10 | 5 | 0.5 |
| L2 | 33 | 15 | 15 |  |
| M | 43 | 11 | 14 |  |
| MS | 5 | 1 | 2 |  |
| **INRG Risk Group** |  |  |  |  |
| Very Low Risk | 1 | 1 | 0 | 0.2 |
| Low Risk | 19 | 10 | 5 |  |
| Intermediate Risk | 31 | 13 | 10 |  |
| High Risk | 48 | 13 | 21 |  |
| **Histology** |  |  |  |  |
| Favorable | 44 | 21 | 13 | 0.2 |
| Unfavorable | 48 | 14 | 20 |  |
| **MYC-N status** |  |  |  |  |
| Negative | 81 | 28 | 33 | 0.09 |
| Positive | 15 | 8 | 2 |  |
| **Loss of CHD5** |  |  |  |  |
| No | 57 | 20 | 24 | 0.4 |
| Yes | 31 | 12 | 9 |  |

| **Resection** |  |  |  |  |
| --- | --- | --- | --- | --- |
| 100% | 30 | 22 | 8 | 0.005 |
| 90 – 99% | 26 | 9 | 17 |  |
| <90% | 17 | 6 | 11 |  |

Age refers to age at diagnosis. INRGSS, International Neuroblastoma Risk Group Staging System. IDRF, Image Defined Risk Factors. VE, vascular encasement.
\* Data in the all patients column may not total 101 due to missing data points: Laterality not available in 1; IDRF not available in 1; INRG Risk Group not available in 2; Histology not available in 9; MYCN data not available in 5; and CHD35 data not available in 13.
\*\* P-values compare the no complication to the complication group for patients who underwent surgery.

### Surgery

In total, 73 patients had a surgical procedure for their tumor. Completeness of resection, as estimated by the surgeon, was 100% in 30, 90% – 99% in 26, and debulking or biopsy only in the remainder. Surgical complications were recorded in 36 of which 4 were Grade I (5%), 19 were Grade II (26%), and 13 were Grade III (18%). There were no grade IV (intensive care unit) or V (death) complications.

### Image Defined Risk Factors

IDRF status was available in 100 patients. One neonate had a resolving left adrenal neuroblastoma diagnosed with a combination of ultrasonography and catecholamines, staged L1, low-risk, and could not be assessed for IDRF for this study because no axial imaging was performed. No IDRF was present in 23 tumors. Of the 77 with one or more IDRF, the median number of IDRF was 3 (IQR 2 – 6, range 1 – 10). One or more vascular encasement IDRF was present in 48; the median number of vascular encasement IDRF in these tumors was 2 (IQR 1 – 3, range 1 – 5).

### Image Defined Risk Factors and complications

The occurrence of a surgical complication correlated with the number of IDRF and with the number of vascular encasement IDRF. In the generalized linear model, complications were significantly associated with IDRF (OR 1.37, 95% CI 1.07 – 1.80, p = 0.01), indicating that the odds of a complication increased 1.37 when the number of IDRF increased by one. Complications correlated more strongly with the subgroup of vascular encasement IDRF (OR 1.86, 95% CI 1.15 – 3.23, p = 0.01) than with the subgroup of non-vascular encasement IDRF (OR 1.35, 95% CI 0.96 – 1.96, p = 0.08).

The cumulative link model showed that the grade of complication did not correlate with the number of IDRF (OR 0.87, 95% CI 0.61 – 1.20, p = 0.4) as demonstrated in Figure 1 which shows a difference in the number of IDRF between complication grade 0 and grade I, but not between grades I, II and III.

**Figure 1.**
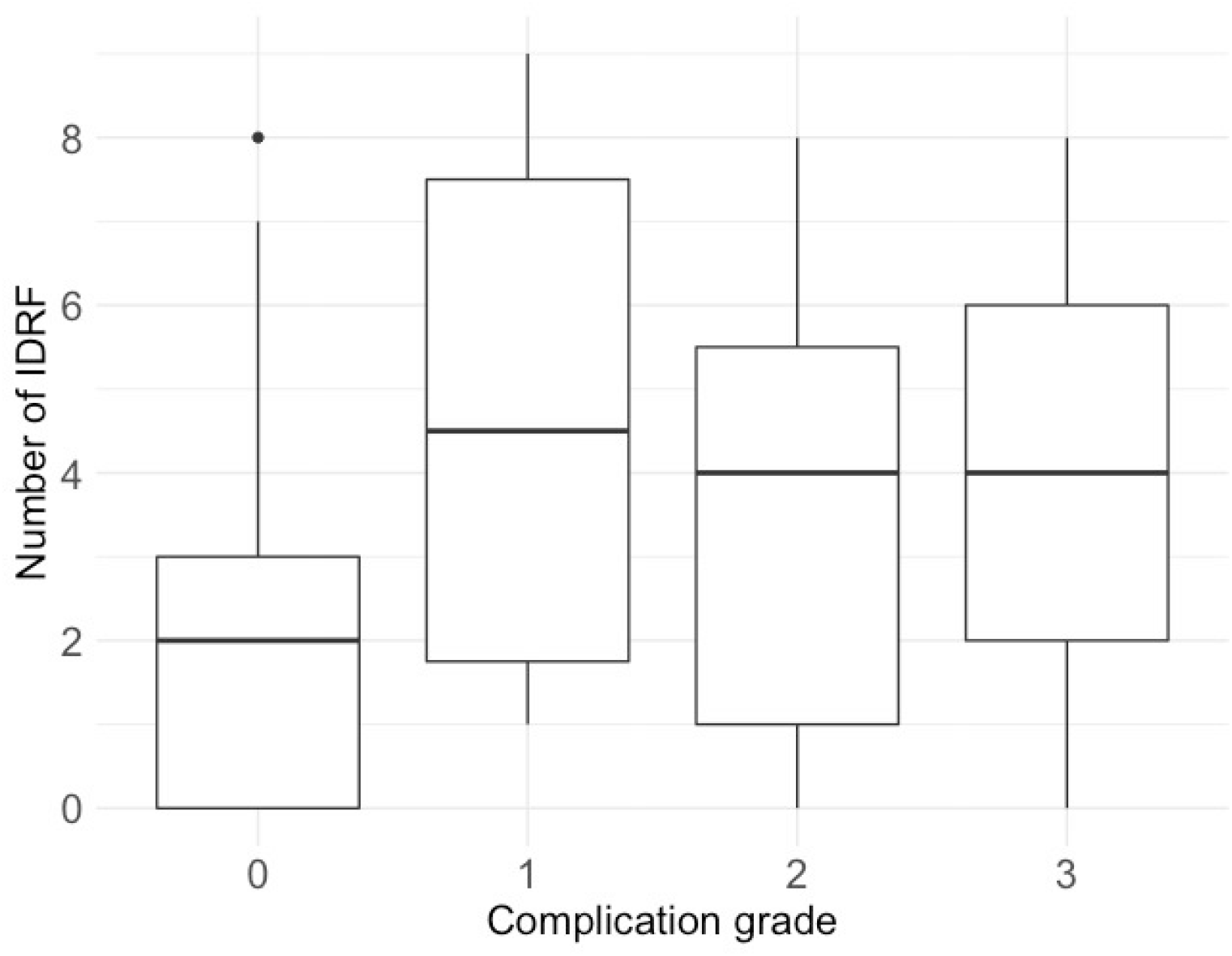
Box plot of the number of Image Defined Risk Factors (IDRF) by Clavien-Dindo grade of complication.

### Survival

In our series, the survival of patients with IDRF was not significantly different from those with no IDRF (Chi square = 0.4, p = 0.5), as shown in Figure 2.

**Figure 2.**
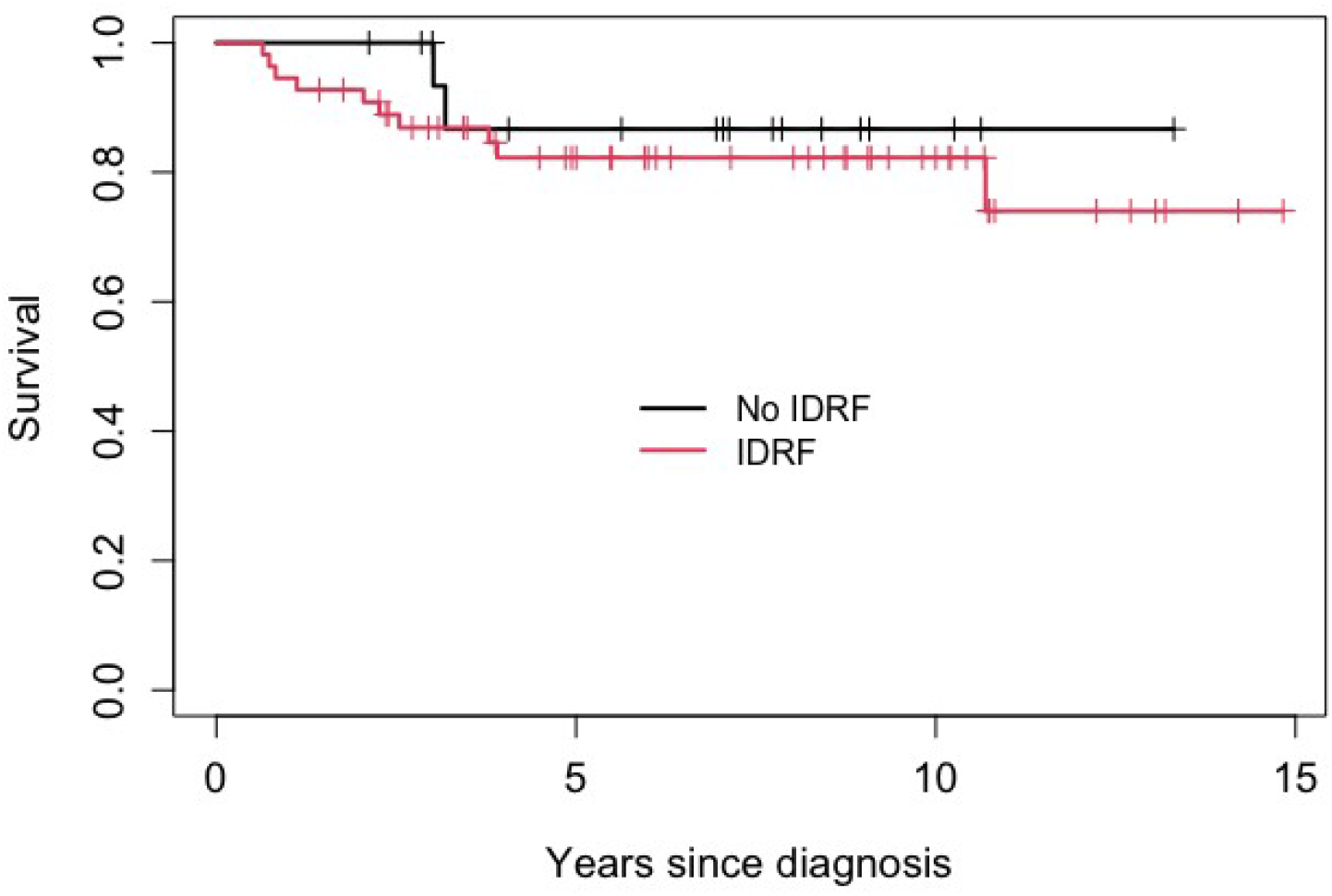
Kaplan-Meyer plot of survival by the presence or absence of IDRF. The difference was not statistically significant (p = 0.5).

### Description of complications

Amongst the complications were four chyle leaks (one thoracic and three abdominal), three hemorrhages secondary to retroperitoneal vessel injury (right gonadal artery, left common iliac vein, left renal vein), two small bowel obstructions requiring adhesiolysis, obstructive jaundice (requiring cholecystostomy tube placement), two nerve injuries (right vocal cord palsy and left phrenic nerve injury), one case of bilateral pneumothoraces, and a transection of the common iliac vein.

All four children with chyle leak received 5 cycles of neoadjuvant chemotherapy. The three patients with abdominal chyle leaks all had encasement of aorta and/or inferior vena cava (IDRF number 16) and encasement of origin of celiac axis or origin of SMA (IDRF number 14). Two of these patients underwent relook laparotomy after failure of medical management by percutaneous drainage and intravenous nutrition. In both children, the chyle leak was identified as coming from lymphatic tissue surrounding the origin of the celiac axis. Both were successfully treated by suture ligation of the leaking lymphatic channels. The third patient’s chyle leak responded to medical management without the need for surgical repair. The chylothorax patient was IDRF positive for infiltration of costovertebral junction T9 – T12 (IDRF number 10) and ipsilateral extension of the tumor within two body compartments (IDRF number 1).

## Discussion

This study shows that IDRF do not all predict surgical complications equally: vascular encasement IDRF predicted complications to a greater extent than other IDRF. IDRF status has been previously shown to correlate with surgical complications [8,18], resectability [7], and survival [19]. Of emerging importance is the association of postoperative complications with specific IDRF groups, for example, those of the vascular encasement type [9] as in our series, those of the organ invasion type [10], or tumor in multiple body compartments [10]. Specific IDRF groups appear to predict specific complications. Lim et al. showed that the risk of nephrectomy was higher with renal vascular involvement or renal invasion [20]. Mansfield et al. reported one case of chyle leak associated with encasement of the celiac axis [6]. Our series adds to this with three further cases of chyle leak associated with the celiac axis and/or superior mesenteric artery encasement IDRF.

One value of the IDRF system is that surgical challenges are translated for the rest of the oncology team and incorporated into staging and risk grouping. We suggest that a more nuanced understanding of specific IDRF could facilitate collaboration between the pediatric surgeon, radiologist, and oncologist. Our findings confirm those of others that not all IDRF are equal [6,7] and that vascular encasement IDRF are associated with an increased risk of complications [9,21]. IDRF status not only correlates with general complications such as hemorrhage, but also with specific complications such as chyle leak. This suggests that, in planning neuroblastoma resection, radiologists and surgeons could work together to highlight specific IDRF that might increase the risk of specific complications and plan steps to mitigate this risk. Radiological reporting of neuroblastomas could highlight specific IDRF, perhaps using a standardized neuroblastoma reporting format along the lines of the recently published International Neuroblastoma Surgical Reporting Form [22]. It will be helpful for less experienced pediatric surgical oncologists to be wary of specific complications associated with certain IDRF and take care to avoid these. From our experience with chyle leaks, we take extra care to ligate and clip lymphatic channels around the celiac and superior mesenteric origin when dissecting neuroblastoma at this site. In the future, newer approaches such as indocyanine green may not only aid neuroblastoma resection [23] but might also be employed intraoperatively to identify and remedy lymphatic leaks at the time of primary resection [24].

### Limitations

This study was limited by its relatively small size. It was a single-center experience. Our complication rate was higher than those published (9 – 28%) [1,8]. This is likely due to the higher proportion of IDRF positive patients in our series and the inclusion of relatively minor complications (Clavien-Dindo I and II [13]). By comparison, in the largest published series to date, 30% had surgical risk factors and complications were reported simply as nonfatal or fatal [1]. Our grade III complication rate was within the range of other published series. Others have shown that IDRF status can change after neoadjuvant chemotherapy [6,7,25,26]; however, our study used IDRF status at diagnosis only. We included one single chromosome aberration marker in our models, loss of CDH5 on chromosome 1p, but had insufficient data to include loss of ATM on chromosome 11q.

## Conclusions

Specific IDRF are of more importance to the surgeon than the presence or absence of any IDRF. Close collaboration between oncologist, radiologist and surgeon is recommended in pre-operative planning of neuroblastoma resection in the presence of IDRF. Future research could take a larger number of chyle leak complications and correlate these patients with pre- and post-neoadjuvant therapy IDRF.

## Supporting information

STROBE

## Data Availability

All data produced in the present study are available upon reasonable request to the authors

